# An Open Repository of Real-Time COVID-19 Indicators

**DOI:** 10.1101/2021.07.12.21259660

**Authors:** Alex Reinhart, Logan Brooks, Maria Jahja, Aaron Rumack, Jingjing Tang, Sumit Agrawal, Wael Al Saeed, Taylor Arnold, Amartya Basu, Jacob Bien, Ángel A. Cabrera, Andrew Chin, Eu Jing Chua, Brian Clark, Sarah Colquhoun, Nat DeFries, David C. Farrow, Jodi Forlizzi, Jed Grabman, Samuel Gratzl, Alden Green, George Haff, Robin Han, Kate Harwood, Addison J. Hu, Raphael Hyde, Sangwon Hyun, Ananya Joshi, Jimi Kim, Andrew Kuznetsov, Wichada La Motte-Kerr, Yeon Jin Lee, Kenneth Lee, Zachary C. Lipton, Michael X. Liu, Lester Mackey, Kathryn Mazaitis, Daniel J. McDonald, Phillip McGuinness, Balasubramanian Narasimhan, Michael P. O’Brien, Natalia L. Oliveira, Pratik Patil, Adam Perer, Collin A. Politsch, Samyak Rajanala, Dawn Rucker, Chris Scott, Nigam H. Shah, Vishnu Shankar, James Sharpnack, Dmitry Shemetov, Noah Simon, Benjamin Y. Smith, Vishakha Srivastava, Shuyi Tan, Robert Tibshirani, Elena Tuzhilina, Ana Karina Van Nortwick, Valérie Ventura, Larry Wasserman, Benjamin Weaver, Jeremy C. Weiss, Spencer Whitman, Kristin Williams, Roni Rosenfeld, Ryan J. Tibshirani

**Affiliations:** Department of Statistics & Data Science, Carnegie Mellon University; Machine Learning Department, Carnegie Mellon University; Computational Biology Department, Carnegie Mellon University; Google.org Fellows, Google LLC; Computer Science Department, Carnegie Mellon University; Linguistics Program, University of Richmond; Information Networking Institute, Carnegie Mellon University; Department of Data Sciences and Operations, University of Southern California; Human-Computer Interaction Institute, Carnegie Mellon University; School of Natural Sciences and Mathematics, University of Texas at Dallas; College of Fine Arts and Department of Psychology, Carnegie Mellon University; Department of Statistics, University of California, Davis; Microsoft Research New England; Department of Statistics, University of British Columbia; Department of Statistics, Stanford University; Department of Biomedical Data Science, Stanford University; Department of Medicine, Stanford University; Program in Immunology, Stanford University School of Medicine; Department of Biostatistics, University of Washington; Heinz College of Information Systems and Public Policy, Carnegie Mellon University

## Abstract

The COVID-19 pandemic presented enormous data challenges in the United States. Policy makers, epidemiological modelers, and health researchers all require up-to-date data on the pandemic and relevant public behavior, ideally at fine spatial and temporal resolution. The COVIDcast API is our attempt to fill this need: operational since April 2020, it provides open access to both traditional public health surveillance signals (cases, deaths, and hospitalizations) and many auxiliary indicators of COVID-19 activity, such as signals extracted from de-identified medical claims data, massive online surveys, cell phone mobility data, and internet search trends. These are available at a fine geographic resolution (mostly at the county level) and are updated daily. The COVIDcast API also tracks all revisions to historical data, allowing modelers to account for the frequent revisions and backfill that are common for many public health data sources. All of the data is available in a common format through the API and accompanying R and Python software packages. This paper describes the data sources and signals, and provides examples demonstrating that the auxiliary signals in the COVIDcast API present information relevant to tracking COVID activity, augmenting traditional public health reporting and empowering research and decision-making.

---

Public health decision makers, healthcare providers, epidemiological researchers, employers, institutions, and the general public benefit from promptly and readily accessible data regarding COVID-19 activity levels, countermeasures, and pandemic impact. Realtime indicators of COVID-19 activity levels, such as statistics on cases, deaths, test positivity, and hospitalizations, enable reports and interactive dashboard applications for situational awareness [1–3], and are essential for most analyses of the pandemic. These data are available for locations across the United States from a number of official sources and independent aggregators in varied and inconsistent formats. Different data types and sources vary in timeliness, based on when measured events occur in the progression of the disease, testing capabilities, the reporting pipeline, and their publication schedules.

Additional, auxiliary data sources can improve on the timeliness, scope, and utility of the “topline” indicators (cases, test positivity, hospitalizations, deaths) coming from the public health reporting system. For example, in the context of other infectious diseases: syndromic surveillance in ambulatory clinics and emergency rooms improves the accuracy of outbreak detection for emerging pathogens such as H1N1 [4]; and digital surveillance (based on, e.g., search and social media trends) enables more accurate “nowcasts” and forecasts of traditional disease surveillance streams such as the CDC’s ILINet [5, 6], as do publication formats providing access to historical versions of a given data set [7, 8]. Several other examples exist that span a wide variety of data platforms and diseases [9–12]. During the COVID-19 pandemic, digital data streams have permitted faster prediction of case increases [13, 14], while enabling analyses of the impact of public health policies on public behavior, the economy, and disease spread [15–18].

The Delphi group worked with partner organizations and public data sets to build a large-scale database of indicators tracking COVID-19 activity and other relevant phenomena in the United States, which has been publicly available and continuously updated since April 2020. Alongside public data on reported cases and deaths, this database includes several unique data streams, including indicators extracted from de-identified medical claims data, antigen test results from a major testing manufacturer, large-scale public surveys that measure symptoms and public behavior, and indicators based on particular Google search queries. (We use the terms “indicator” and “signal” interchangeably.) We make aggregate signals publicly available, generally at the county level, via the COVIDcast API [19]. We store and provide access to all previous (historical) versions of the signals, a key feature that exposes the effects of data revisions. Moreover, we provide R [20] and Python [21] packages to facilitate interaction with the API, and an online dashboard to visualize the data [22].

In a companion paper, we analyze the utility provided by a core set of the indicators in short-term COVID-19 forecasting and hotspot prediction models [23]. In another companion paper, we elaborate on our research group’s (Delphi’s) large-scale public surveys, run in partnership with Facebook and available in aggregate form in the COVIDcast API [24]. This paper focuses on the COVID-19 indicators themselves, describing the data streams, how they are processed and made publicly available, and insights that can be gained by combining novel data sources with standard public health surveillance data.

## 1 Methods

### 1.1 Data Collection

We receive data daily from healthcare partners, technology companies, and from surveys conducted daily by Delphi in partnership with Facebook. These data sources provide information not available from standard public health reporting or other common sources, such as:

#### Health Insurance Claims

Based on de-identified medical insurance claims from Change Healthcare and other health system partners, we release indicators on the estimated percentage of covered outpatient visits and hospitalizations that involved COVID diagnoses or symptoms.

#### Internet-Based Surveys

Conducted in partnership with Facebook, Delphi’s COVID-19 Trends and Impact Survey receives an average of 50,000 responses daily, and has received over 25 million responses since April 2020 [24, 25]. From the surveys, we construct indicators on symptoms, social distancing, vaccination, and other attitudes and behaviors related to COVID. The surveys are voluntary and participants are redirected to a platform managed by Carnegie Mellon University to give consent and take the survey; individual response data is not provided to Facebook and the data collection protocol was approved by the Carnegie Mellon Institutional Review Board (STUDY2020_00000162).

#### COVID Antigen Tests

Based on data from Quidel, a manufacturer of COVID antigen tests in the United States, we calculate and release (Quidel-specific) test volumes and positivity rates.

#### Search Trends

Based on Google’s COVID-19 Search Trends data set [26], we provide indicators reflecting COVID-related search activity.

#### Mobility Data

SafeGraph, a company that collects geospatial data from smartphone apps, calculates COVID-related mobility signals [27, 28] and makes them available to researchers under a data use agreement; we aggregate (some of) these signals to the county level and make them publicly available.

We also scrape data accessible from other public sources, such as cases and deaths data aggregated from public reporting by JHU CSSE [1] and by USAFacts [3], so that we can track revisions and updates to this data (see Section 1.3).

Altogether, we produce over 170 signals from 12 distinct sources, and provide them in a common format for access. This unifies both unique (unavailable anywhere else) and standard COVID data streams into a single common format, enabling efficient comparison and modeling. A summary of the data sources and signals in the API is in Table 1, and detailed documentation is available online at https://cmu-delphi.github.io/delphi-epidata/api/covidcast_signals.html.

**Table 1:** Data sources available in the COVIDcast API [19], as of date of publication. The first group of data sources are produced from data not otherwise available publicly (or only available in limited form); the second group is mirrored from public sources.

| Data source | Signals available | First date | Resolution |
| --- | --- | --- | --- |
| Change Healthcare | Percentage of outpatient visits with COVID diagnostic codes or codes indicating COVID-like symptoms. Based on de-identified claims data processed by Change Healthcare. | 2020-02-01 | County* |
| Doctor Visits | Percentage of outpatient visits primarily about COVID-like symptoms, based on de-identified claims data provided by health system partners. | 2020-02-01 | County* |
| Hospital Admissions | Percentage of new hospital admissions with COVID diagnostic codes, based on de-identified claims data provided by health system partners. | 2020-02-01 | County** |
| Quidel | Test positivity rates for COVID-19 antigen tests produced by Quidel. | 2020-05-26 | County** |
| SafeGraph | Mobility metrics, such as time away from home or visits to bars and restaurants, based on cell phone mobility data collected by SafeGraph [27, 28]. | 2019-01-01 | County |
| COVID-19 Trends and Impact Survey | COVID symptoms, social distancing behaviors, mental health, economic impact, behavior (e.g., mask-wearing, vaccination attitudes), and COVID testing signals based on daily surveys conducted nationally by Delphi through Facebook [24, 25]. | 2020-04-06 | County** |
| Health & Human Services | Counts of hospital admissions due to confirmed or suspected COVID-19, as reported by the Department of Health & Human Services. | 2019-12-31 | State |
| COVID Act Now | COVID-19 testing results, such as positivity rate and number of tests, compiled by COVID Act Now from CDC reporting. | 2020-03-02 | County* |
| Google Symptoms | Trends in Google search volume for terms related to anosmia and ageusia (loss of smell or taste), which correlate with COVID activity, based on data shared by Google [26]. | 2020-02-13 | County*** |
| Cases and Deaths | Confirmed COVID-19 cases and deaths, compiled by JHU CSSE [1] and by USAFacts [3]. | 2020-01-22 | County |
| NCHS Mortality | Weekly totals of deaths broken down by cause, such as COVID, flu, or pneumonia, compiled by the National Center for Health Statistics [29]. | 2020-01-26 | State |

\*Available at > 60% of counties; \*\*available at 20–60% of counties; \*\*\*available at < 20% of counties. For some signals, location availability varies over time, e.g., due to variable reporting volume.

### 1.2 Signal Processing

Because each data source reports data in different formats, we must convert each source to a common format. In this format, each record represents an observation of one quantity at one time point in one location. Locations are coded consistently using standard identifiers such as FIPS codes; the sample size and standard error for each observation is also reported when applicable. Each signal is reported at the finest geographic resolution its source supports (such as county or state) and also aggregated to metropolitan statistical areas, Health and Human Services regions, and hospital referral regions. National averages are also provided. Crucially, each record is tagged with an *issue date* referring to when the value was first issued, as described below. This allows tracking of revisions made to individual observations, as each revision is tagged with its own issue date.

When appropriate, additional post-processing (often nontrivial) is applied to the data. For example, data on visits to doctors’ offices is subject to strong day-of-week effects, and so regression is used to adjust for these effects. Other indicators are available in raw versions and versions smoothed with a 7-day trailing average. All processing is done using open-source code written primarily in Python and R, and available publicly at https://github.com/cmu-delphi/covidcast-indicators/. The processing steps used for each signal are publicly documented on their respective pages at https://cmu-delphi.github.io/delphi-epidata/api/covidcast_signals.html.

### 1.3 Revision Tracking

Many data sources that are useful for epidemic tracking are subject to revision after their initial publication. For example, aggregated medical claims data may be initially published after several days, but additional claims and corrections may take days to weeks to be discovered, processed, and aggregated. Medical testing data are also often subject to backlogs and reporting delays, and estimates for any particular date are revised over time as errors are found or additional data becomes available. This revision process is generally referred to as *backfill*.

For this reason, the COVIDcast API annotates every observation with two dates: the *time value*, the date the underlying events (such as tests or doctor’s visits) occurred, and the *issue date* when we aggregated and reported the data for that time value. Importantly, there can be multiple observations for a single time value with different issue dates, for example if data is revised or claims records arrive late. We track revisions to all data sources included in the API, including external data sources (such as sources tracking cases and deaths). Many external sources do not keep a public or conveniently accessible record of revisions of their data.

For many purposes it is sufficient to use the most recently issued observation at a given time value, and the COVIDcast API returns the most recent issue as its default. However, for some applications it is crucial to know what was known *as of* a specific date. For example, an epidemic forecasting model will be called upon to make its forecasts based on preliminary data about recent trends, so when it is trained using historical data, it should be trained using the initial versions of that data, not updates that would have been received later. Moreover, these revision records allow models to be modified to account for noise and bias in early data versions, or to exclude data that is too new to be considered stable, and to “rewind” time and simulate how these revised models would have performed using only the versions of data available *as of* those times.

Research on data revisions in the context of influenza-like illness has shown that backfill can significantly alter forecast performance [7, 30], and that careful training on preliminary data can reduce this influence [8]. Recent research has shown similar results for COVID-19 forecasts [31]. We also examine this in our companion paper on forecasting, where we observe that training and validating models on finalized data yields overly optimistic estimates of true test-time performance [23].

### 1.4 Public API

The data described above is publicly available through the Delphi COVIDcast API [19]. By making HTTP requests specifying the data source, signal, geographic level, and time period desired, users can receive data in JSON or CSV form. For added convenience, we have written covidcast R [20] and Python [21] packages with functions to request data, format it as a data frame, plot and map it, and combine it with data from other sources.

The R and Python package software is public and open-source, at https://github.com/cmu-delphi/covidcast/. The API server software is itself also public and open-source, at https://github.com/cmu-delphi/delphi-epidata/. Lastly, most data sources are provided under the Creative Commons Attribution license, and a small number have additional restrictions imposed by the data source; see https://cmu-delphi.github.io/delphi-epidata/api/covidcast_licensing.html.

### 1.5 Interactive Visualization

Since April 2020, we have been maintaining and continually improving various online visualization tools for the COVIDcast indicators [22]. These tools fetch data directly from the API, and allow for exploration of both temporal (e.g., time series graphs) and spatial (e.g., choropleth maps) trends in the signals, as well as many other aspects, such as correlations, anomalies, and backfill. There is also a dedicated dashboard for exploring results from the COVID-19 Trends and Impact Survey. The visualizations have been continually improved as new sources of data arrive, and in response to interviews with users and health experts, usage analytics from the site, and user surveys.

### 1.6 COVID Forecasting

Since July 2020, we have been regularly submitting short-term forecasts of COVID-19 case and death incidence, at both the state and county levels, to the COVID-19 Forecast Hub [32], with “CMU-TimeSeries” as the team-model name. The process of building, training, and deploying our forecasting models leverages much of the infrastructure described in this paper (such as the COVIDcast API’s *as of* feature), and some of our forecasting systems rely on auxiliary indicators (such as survey-based and claims-based COVID-like illness signals, which are described below).

## 2 Results

The indicators that are available in the COVIDcast API have been used in dashboards produced by COVID Act Now [33], COVID Exit Strategy [34], and others; to inform the Delphi, DeepCOVID [35], and the Institute for Health Metrics and Evaluation (IHME) [36] COVID forecasting models; in various federal and state government reports and analyses; and in a range of news stories. Aside from operational use in decision-making and forecasting, they have also facilitated numerous analyses studying the impacts of COVID-19 on the public, the effectiveness of policy interventions, and factors that influenced the spread of the pandemic [17, 18, 37–40]. The API currently serves hundreds of thousands of requests to thousands of users every day.

In what follows, we present examples of the usefulness of some of the novel signals available in the API. These examples demonstrate that such indicators are meaningfully related to COVID activity, that they provide alternate views on pandemic activity that are not subject to the same reporting glitches and delays as traditional public health surveillance streams, and that they provide information about public behavior and attitudes that are not available from any other source. Code to reproduce all examples (which uses the covidcast R package and fetches data from the API) can be found at https://doi.org/10.5281/zenodo.5639567.

### 2.1 Tracking Trends

Many of the indicators in the COVIDcast API are intended to track COVID activity. Five indicators in particular have the closest connections to confirmed cases:

- Change Healthcare COVID-like illness (CHNG-CLI): The percentage of outpatient visits that are primarily about COVID-related symptoms, based on de-identified Change Healthcare claims data.
- Change Healthcare COVID (CHNG-COVID): The percentage of outpatient visits with confirmed COVID-19, based on the same claims data.
- COVID-19 Trends and Impact Survey CLI (CTIS-CLI): The estimated percentage of the population with COVID-like illness based on Delphi’s surveys of Facebook users.
- COVID-19 Trends and Impact Survey CLI in the community (CTIS-CLI-in-community): The estimated percentage of the population who know someone in their local community who is sick, based on the same surveys.
- Quidel test positivity rate (Quidel-TPR): The percentage of positive results among Quidel COVID antigen tests.

Figure 1 compares the first three of these signals to COVID cases in the United States (from JHU CSSE, smoothed with a 7-day trailing average) over a year of the pandemic (April 15, 2020 to April 15, 2021), illustrating how they track national trends quite well. Importantly, this same relationship persists across multiple resolutions of the data, down to smaller geographic regions such as states and counties, as shown in the Supplementary Information. This will also be illustrated in a more detailed correlation analysis in the next subsection.

**Figure 1:**
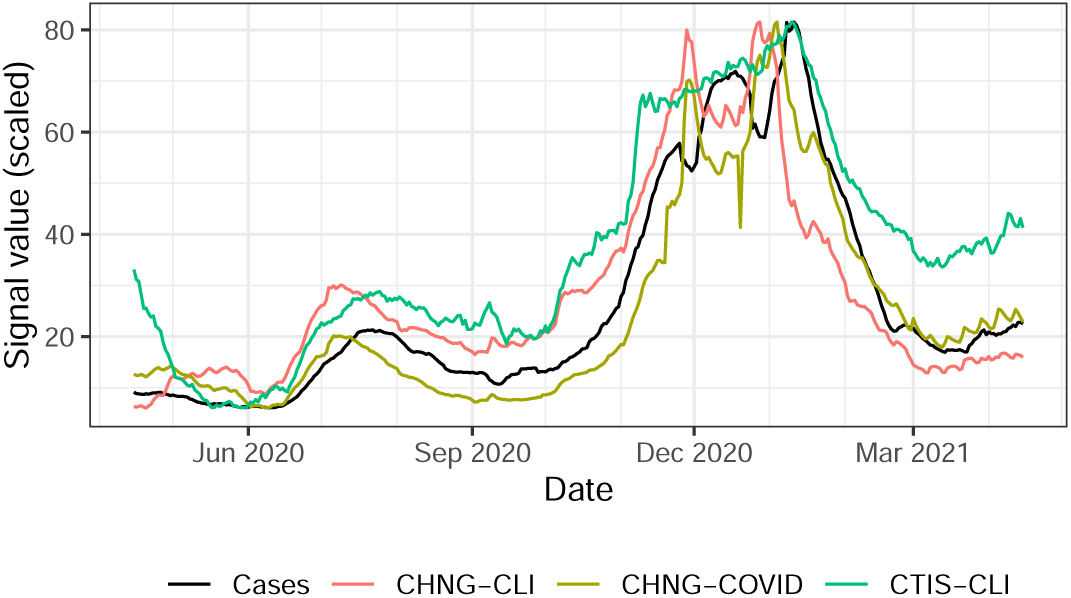
National trends, from April 2020 to April 2021, of four signals in the COVIDcast API. The auxiliary signals, based on medical claims data and massive surveys, track changes in officially reported cases quite well. (They have all been placed on the same scale as reported cases per 100,000 people.)

Besides tracking contemporaneous COVID activity, these and other indicators can be used to improve forecasts of future COVID case trends, as investigated in our companion paper [23].

### 2.2 Correlation Analyses

To quantify the ability of the signals described above to track trends in COVID cases, we use the Spearman (rank) correlation and analyze two key correlation patterns, between each signal and confirmed COVID case rates (cases per 100,000 people):

1. *Geo-wise correlations* (i.e., on a specific date, do values of the signal correlate with case rates across locations?): Formally, let *X*_*t*_ and *Y*_*t*_ be vectors of values of a signal and case rates, over all locations, on date *t*. The geo-wise correlation at time *t* is defined as cor(*X*_*t*_, *Y*_*t*_) (where here and throughout cor(·,·) denotes Spearman correlation). This examines whether a signal has the capability to help spot locations with high case rates at any given time.
2. *Time-wise correlations* (i.e., at a specific location, do values of the signal correlate with case rates across time?): Let *X*_*ℓ*_ and *Y*_*ℓ*_ be vectors of values of a signal and case rates, over all times, at location *ℓ*. The time-wise correlation at location *ℓ* is defined as cor(*X*_*ℓ*_, *Y*_*ℓ*_). This examines whether changes in a signal over time correspond to changes in reported cases at the same location.

Figure 2 shows the geo-wise correlations achieved by the five signals and COVID case rates (from JHU CSSE, smoothed using a 7-day trailing average), from April 15, 2020 to April 15, 2021. This calculation is performed over all counties with at least 500 cumulative cases by the end of this period, and at which all indicators are available (956 counties in total). The large positive correlations suggest that these signals could be useful in hotspot detection (identifying counties that have relatively high COVID activity, at a given time). Somewhat surprisingly, the survey-based CLI-in-community signal shows the strongest correlations for much of the time period. This clearly demonstrates the value of a large-scale survey such as CTIS for tracking symptoms and case trends, especially when other data is unavailable.

**Figure 2:**
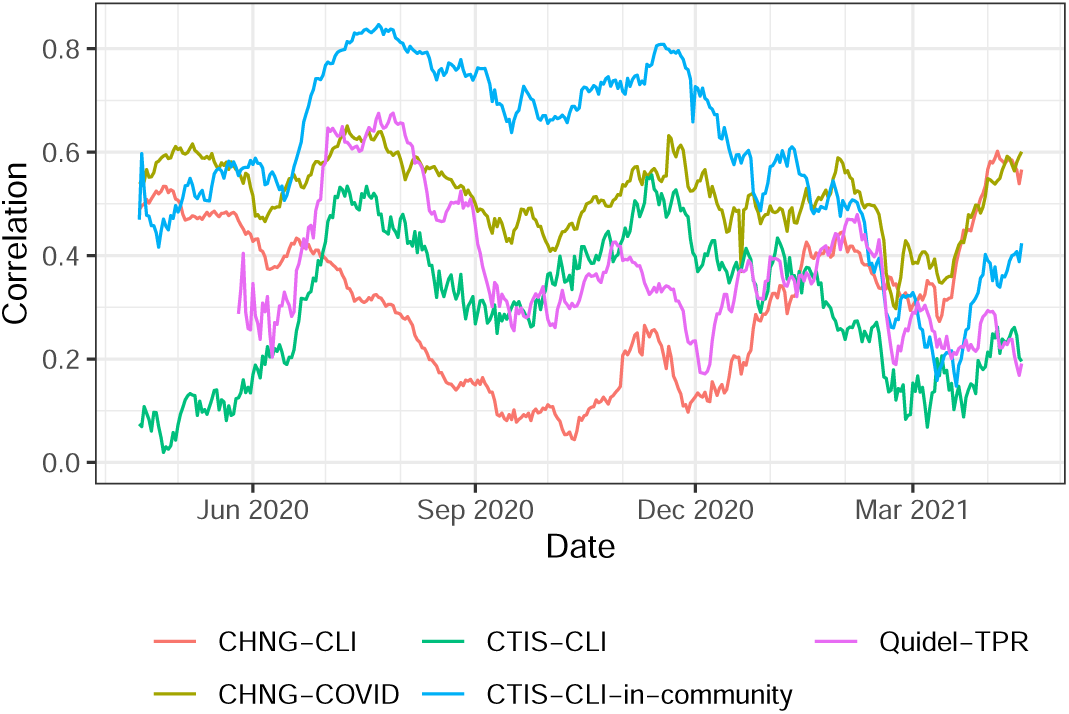
Geo-wise correlations with case rates, from April 15, 2020 to April 15, 2021, calculated over all counties for which all signals were available and which had at least 500 cumulative cases by the end of this period.

Also notable is the fact that the correlations fluctuate over time in complex ways; while some of this variation is likely due to changes in public behavior, reporting and testing practices, and so on, some is also related to changes in overall COVID-19 case trends. For example, comparing against Figure 1, we see that correlations drop in February 2021 for many of the signals, roughly matching the point in time when COVID cases also sharply decline. This decline likely caused the heterogeneity in case rates by county to decline, making it more difficult for any signal to achieve a high geo-wise correlation. Correlations between confirmed cases and cases 1–3 weeks prior (shown in the Supplementary Information) show a similar correlation drop in February 2021, showing that the drop is due to a change in case data and not problems with the other signals.

Figure 3 summarizes time-wise correlations from these five signals over the same time period, and for the same set of counties. For each signal, we display the set of correlations that it achieves in histogram form (more precisely, using a kernel density estimate). All signals produce positive correlations in the majority of counties considered (with very little mass in each estimated density being to the left of zero). The largest correlations, in bulk, are achieved by the CHNG-COVID signal; the CTIS-CLI-in-community signal is a close second, and the CHNG-CLI signal is third. There are two noteworthy points:

**Figure 3:**
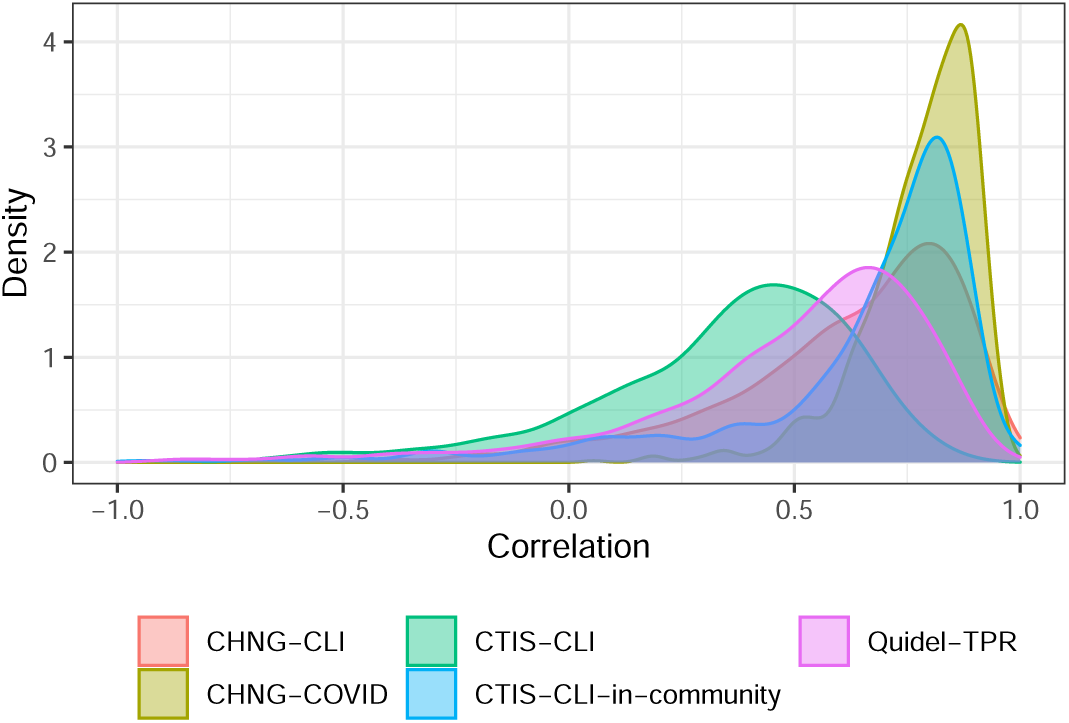
Time-wise correlations with case rates, from April 15, 2020 to April 15, 2021, calculated over all counties for which all signals were available and which had at least 500 cumulative cases by the end of this period.

- This is different from what is observed in Figure 2, where the CTIS-CLI-in-community signal achieves clearly the highest correlations for most of the time period. However, it is worth emphasizing that time-wise and geo-wise correlations are truly measuring different properties of a signal; and the claims signals (CHNG-COVID and CHNG-CLI) seem more appropriate for temporal—rather than spatial—comparisons. We revisit this point in the discussion.
- It is still quite impressive (and surprising) that the CTIS-CLI-in-community signal, based on people reporting on the symptoms of others around them, can achieve nearly as strong time-wise correlations to confirmed cases as can a signal that is based on picking up the occurrence of a confirmed case passing through the outpatient system.

The Supplementary Information contains additional correlation analyses that compare these COVID-related signals to COVID hospitalizations reported by the Department of Health and Human Services. These show similar results, illustrating that the signals are useful for tracking key health outcomes.

### 2.3 Helping Robustness

Public health reporting of COVID tests, cases, deaths, and hospitalizations is subject to a number of possible delays and problems. For example, COVID testing data is reported inconsistently by different states using different definitions and inclusion criteria, and differences in reporting processes mean state data often does not match data reported to the federal government [41]. Case and death data is frequently backlogged and corrected, resulting in artificial spikes and drops [42, 43].

As an example, looking back at Figure 1, we can see clear dips in the confirmed COVID case curve that occur around the Thanksgiving and New Year’s holidays. This is artificial, and due to the fact that public health departments usually close over holiday periods, which delays case and death reporting (for this reason, the artificial dips persist at the state- and county-level as well). This delay denies public health officials timely signals of current trends for the duration of the holidays. The CLI signal from the survey, on the other hand, displays no such dips. The claims signals actually display holiday effects going in the other direction: they exhibit *spikes* around Thanksgiving and New Year’s. This is because they measure the fraction of all outpatient visits with a certain condition, and the denominator (total outpatient visits) drops disproportionately during holiday periods, as people are likely less willing to go to the doctor for more routine issues. Fortunately, in principle, the holiday effects in claims signals should be correctable: they are mainly due to *overall* changes in medical seeking behavior during holiday, periods, and we can estimate such effects using historical claims data.

As a further example, Figure 4 displays data from Bexar County, Texas (which contains San Antonio) during July 2020. On July 16, 2020, San Antonio reported 4,810 backlogged cases after reporting problems prevented them from being reported over the past two weeks [44], resulting in a clearly visible spike in the left-hand panel of the figure (case data from JHU CSSE, smoothed using a 7-day trailing average). Meanwhile, Delphi’s COVID Trends and Impact Survey averaged around 350 responses per day in Bexar County over the same time period, and was able to estimate the fraction of the population who know someone in their local community with COVID-Like Illness (CLI). As we can see in the right-hand panel of the figure, this indicator was not affected by Bexar County’s reporting problems and, as shown in the last subsection, it is (in general) highly correlated with case rates, providing an alternate stream of data about COVID activity unaffected by backlogs. In general, reporting problems have occurred in many jurisdictions across the United States, and audits have regularly discovered misclassified or unreported cases and deaths, making it valuable to cross-check against external sources not part of the same reporting systems.

**Figure 4:**
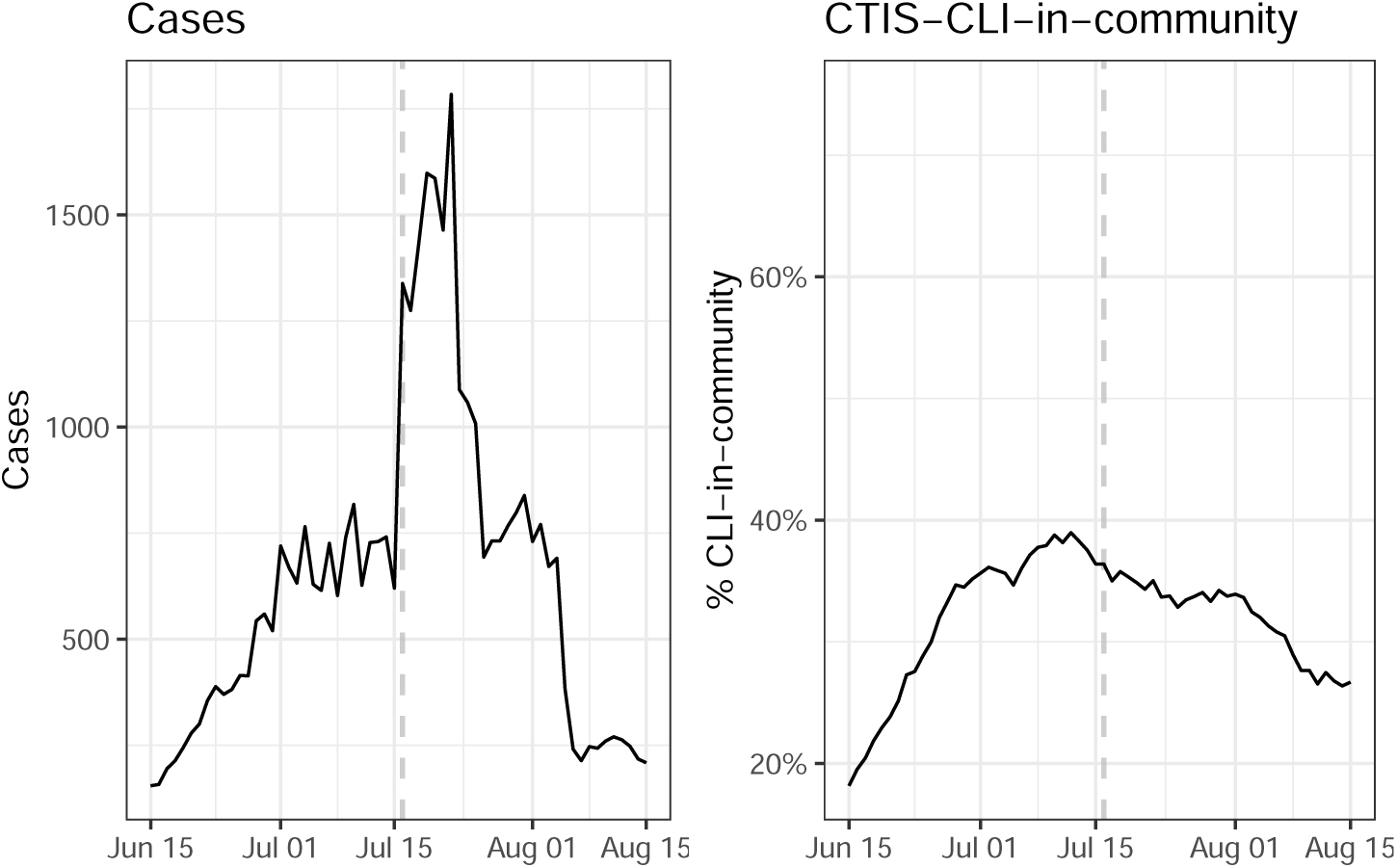
Reported cases per day in Bexar County, Texas during the summer of 2020. On July 16, 4,810 backlogged cases were reported, though they actually occurred over the preceding two weeks (this shows up as a prolonged spike in the left panel due to the 7-day trailing averaging applied to the case counts). Daily CTIS estimates of CLI-in-community showed more stable underlying trends.

### 2.4 Revisions Matter

The revision tracking feature in the API assists in model-building and evaluation. Figure 5 illustrates how DV-CLI, a medical claims signal, evolved as it was revised across multiple issue dates, in four different states, between June 1 and August 1, 2020. DV-CLI is similar to CHNG-CLI and reflects outpatient visits with COVID-related symptoms (the two signals are based on claims data provided by different data partners, which cover different hospital systems). In each panel, the rightmost end of each colored line corresponds to an estimate for the last day of available data for a given issue date, which we can see tends to be significantly biased upward in Arizona in June 2020, and significantly biased downward in New York throughout June and July 2020.

**Figure 5:**
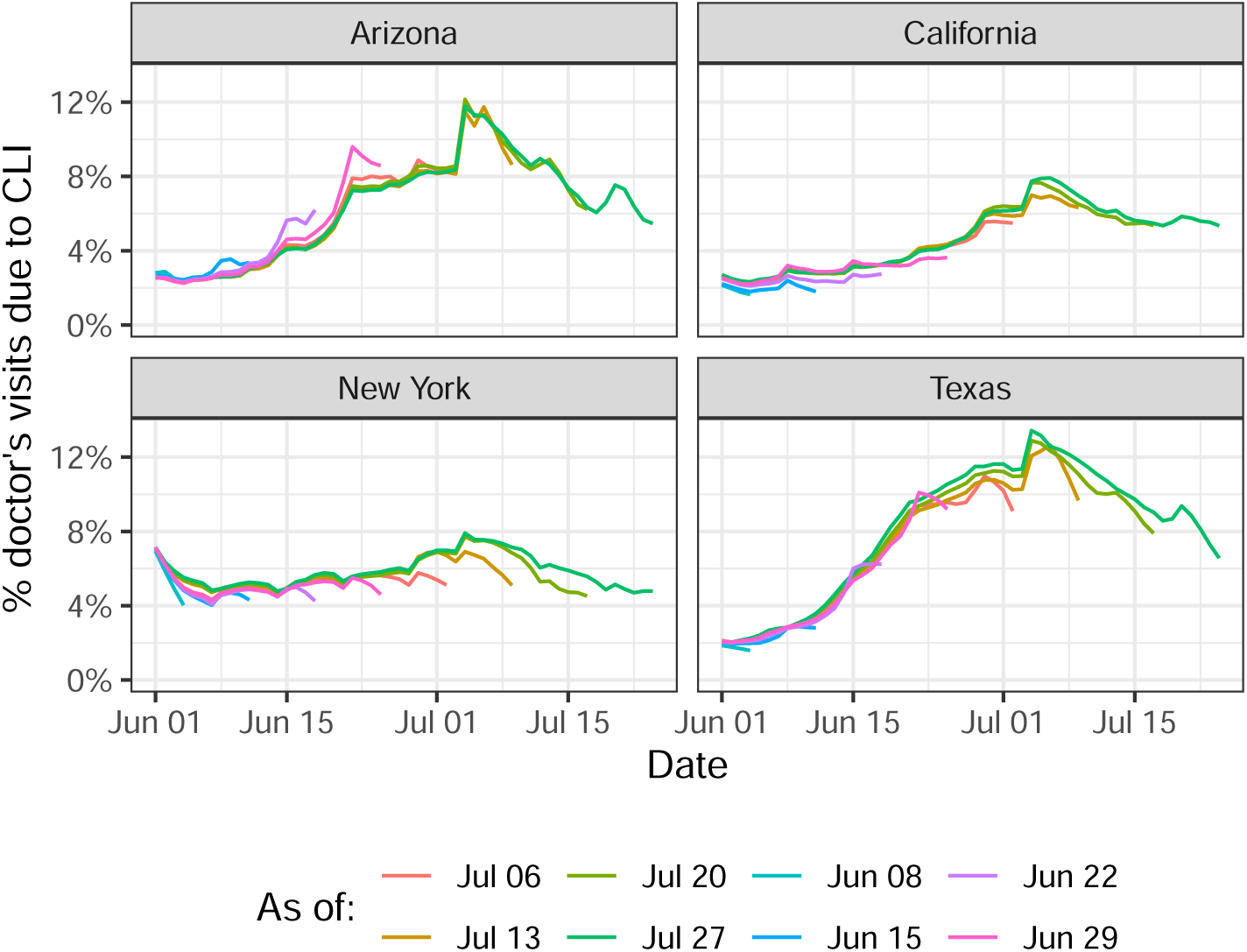
Estimated percentage of outpatient visits due to COVID-like illness (DV-CLI) displayed across multiple issue dates, with later issue dates adding additional data and revising past data from prior issue dates.

Claims-based signals typically undergo heavy backfill as additional claims are processed and errors are corrected; the median relative error between initial reports and final values is over 10% for such data, and only after roughly 30 days do estimates typically match finalized values within 5%. However, the systematic nature of this backfill, as illustrated in Figure 5, suggests that statistical models could be fit (potentially separately for each location) to estimate the final values from preliminary reports.

Figure 6 shows relative differences between early indicator values, reported 10–90 days after the underlying events, and later versions at least four months later. As the distribution of these revision amounts is highly skewed, the figure plots the 95th percentile of relative change, showing that reported deaths can incur large relative error in initial values, comparable to that in claims-based signals. However, for deaths, as well as cases, these large revisions are not very systematic, with large corrections typically occurring at a sparse subset of locations and times (e.g., due to audits or backlogs being cleared, which can result in thousands of cases or deaths can be added or removed all at once). This backfill is much more difficult to predict than that of claims data, therefore the latter (and other sources) may be useful for nowcasting cases and deaths while public health reports are being aggregated and corrected.

**Figure 6:**
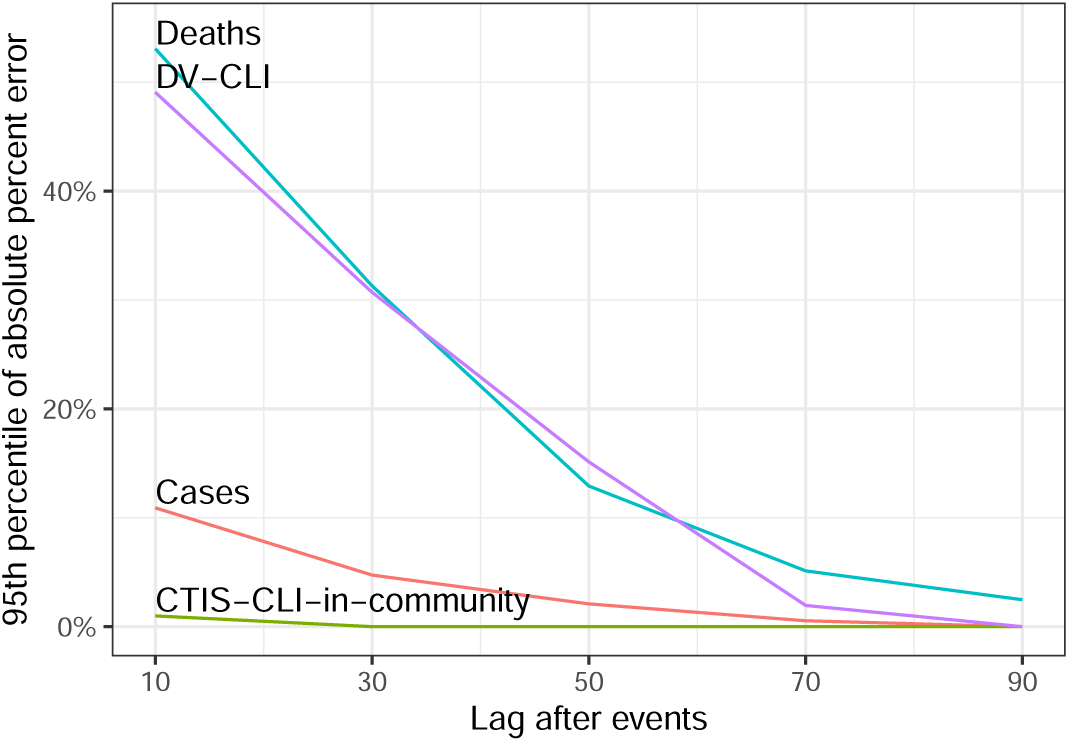
The 95th percentiles of relative error of early reported values of key signals compared to final values reported much later. For each date between October 15, 2020 and April 15, 2021, the values for each state reported between 10 and 90 days later are compared to “final” versions recorded as of August 13, 2021. Even officially reported case and death data can have large revisions 30–60 days or more after initial reporting; much of this is driven by individual large revisions affecting specific states and dates, rather than systematic changes affecting all states and dates.

To reiterate a previous point, when building forecast models (on historical data) for retrospective evaluation purposes, users will want to use data that was known *as of* the forecast date, not revised versions that only became available at a later time. Note that not only model training, but also model assessment, can be affected by the revision process (comparisons of forecasts to the ground truth may shift when the ground truth is revised weeks later). Only by systematically tracking revisions can all these effects be monitored and properly accounted for. The COVIDcast API makes all historical versions available and easily accessible for this purpose; and this feature plays a prominent role in our own analysis of forecasting and hotspot prediction models appearing in a companion paper [23].

### 2.5 New Perspectives

Auxiliary signals (outside of the standard public health reporting streams) can serve as indicators of COVID activity, but they can also illustrate the effect of mitigating actions (such as shelter-in-place orders) and can guide resource allocation for fighting the pandemic. For example, medical claims data reflects healthcare-seeking behavior; measures of mobility reflect adherence to public health recommendations; and measures of COVID vaccine acceptance can guide outreach efforts.

As an illustration, Figure 7 illustrates how CTIS results in January 2021, based on an item asking survey respondents whether they would accept a COVID-19 vaccine if one were offered today, predicted actual uptake of COVID-19 vaccines by July 2021 (as reported by the Centers for Disease Control and Prevention). It also reveals a geographical disparity: in the Northeast, actual vaccination rates more closely match vaccine willingness rates in January than they do in the South, where vaccination rates lag overall. While these results must be interpreted carefully due to potential sampling biases in the survey, they illustrate the potential of data in the COVIDcast API to inform public health decision-making.

**Figure 7:**
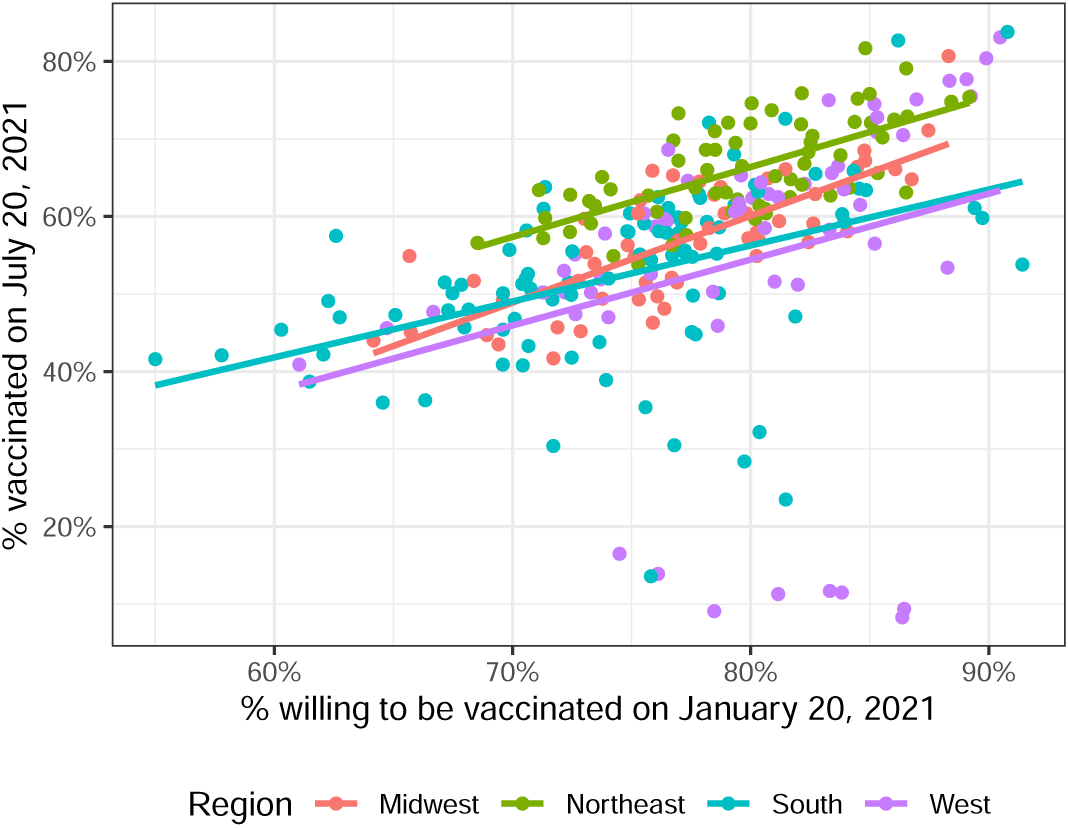
CTIS estimates of the percentage of people willing to get vaccinated, back on January 20, 2021, compared to CDC reporting of the percentage of people vaccinated, on July 20, 2021. Each point is a county (with at least 250 survey responses between January 14–20, 2021), colored by its parent United States Census region.

## 3 Discussion

The COVIDcast API provides open access to real-time and geographically-detailed indicators of COVID activity in the United States, which supports and enhances standard public health reporting streams in several ways.

First, several signals in the API closely track COVID activity (over both time and space); yet they are derived from different data streams (such as surveys, medical insurance claims, and medical devices), and are thus not subject to the same sources of error as public health reporting streams. This can be important both for robustness and situational awareness, allowing decision-makers to diagnose potential anomalies in standard surveillance streams, and for modeling tasks such as forecasting and nowcasting. Our companion paper on forecasting discusses this in more detail [23].

Second, the API features many other signals that are relevant to understanding aspects of the pandemic and its effects on the United States population that are not found in traditional public health streams, such as data on mobility patterns, internet search trends, mask wearing, and vaccine hesitancy, to name just a few. (The latter two signals are derived from the COVID-19 Trends and Impact Survey; our companion paper on this survey gives a more detailed view of its features and capabilities [24].) These signals have already supported pandemic research and policy-making.

Third, the underlying database tracks all revisions made to the data, allowing us to query the API to learn “what was known when,” which is critical for understanding the behavior (and potential pitfalls) of real-time surveillance signals. Such revision data is rarely available in standardized format from other sources.

Finally, we emphasize that unifying many relevant signals into a single common format, with comprehensive revision tracking, is an important goal in and of itself. The ability to combine public health reporting data, syndromic surveillance data, and digital measures of mobility and behaviors goes beyond providing traditional situational awareness. Convenient and real-time access to this data enables continuous telemetry summarizing how things are, how they are expected to change, which areas need additional resources to be allocated in response, and how effective public communication is.

There are a number of open questions, and challenges that remain. Several signals are subject to biases, such as survey sampling and nonresponse biases, geographic differences in market share for medical claims data, or biases in the population represented in app-based mobility data. Claims data tends also to be subject to biases during major national holidays and other events that change healthcare-seeking behavior. Characterizing these biases will be important for future research and operational systems that use these signals. Several data sources are also subject to extensive revision and backfill, which must be studied and modeled to enable effective real-time use of these sources in forecasting and nowcasting systems. The breadth and unique features of the COVIDcast API will help facilitate this and other related work, which will be vital to advancing pandemic modeling and preparedness.

## Data Availability

All data used in the article is freely available in the public COVIDcast API. Code used to produce figures and results is available at https://doi.org/10.5281/zenodo.5639567

https://cmu-delphi.github.io/delphi-epidata/api/covidcast.html

https://doi.org/10.5281/zenodo.5639567

## Acknowledgments

We thank Carrie Reed, Matt Biggerstaff, Michael Johansson, Rachel Slayton, Velma Lopez, Jo Walker and others on the CDC COVID-19 Modeling Team; Hal Varian, Brett Slatkin, and others on Google’s Surveys team; Erin Hattersley, Rasmi Elasmar, and others at Google.org; Evgeniy Gabrilovich and others on Google’s Health team; Curtiss Cobb and others on Facebook’s Demography and Survey Science, Data for Good and Health teams; Alex Smola and others at Amazon Web Services; Tim Suther and others at Change Healthcare; Paul Nielsen and others at Optum; John Tamerius and others at Quidel; and Ross Epstein and others at SafeGraph. This material is based on work supported by gifts from Facebook, Google.org, the McCune Foundation, and Optum; Centers for Disease Control and Prevention (CDC) grant U01IP001121; National Science Foundation Graduate Research Fellowship Program (NSF GRFP) award DGE1745016; and the Center for Machine Learning and Health (CMLH) at Carnegie Mellon.

## Supplemental figures

**Figure 8:**
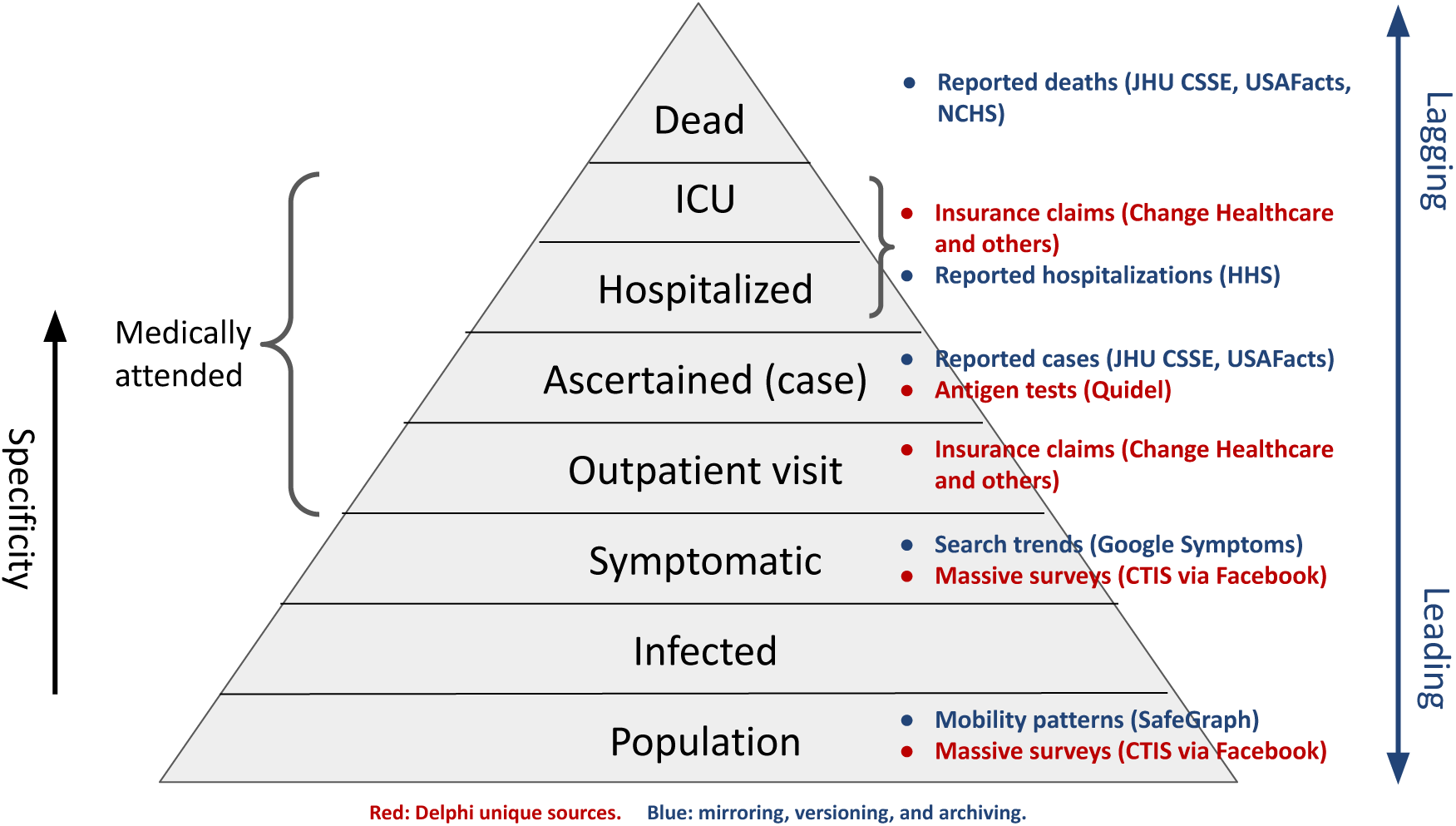
Epidemiological “severity pyramid”, representing the progression of disease progression, from relevant public behaviors, through infection, towards increasingly severe stages of disease. The annotations here refer to the data sources available in Delphi’s Epidata API.

**Figure 9:**
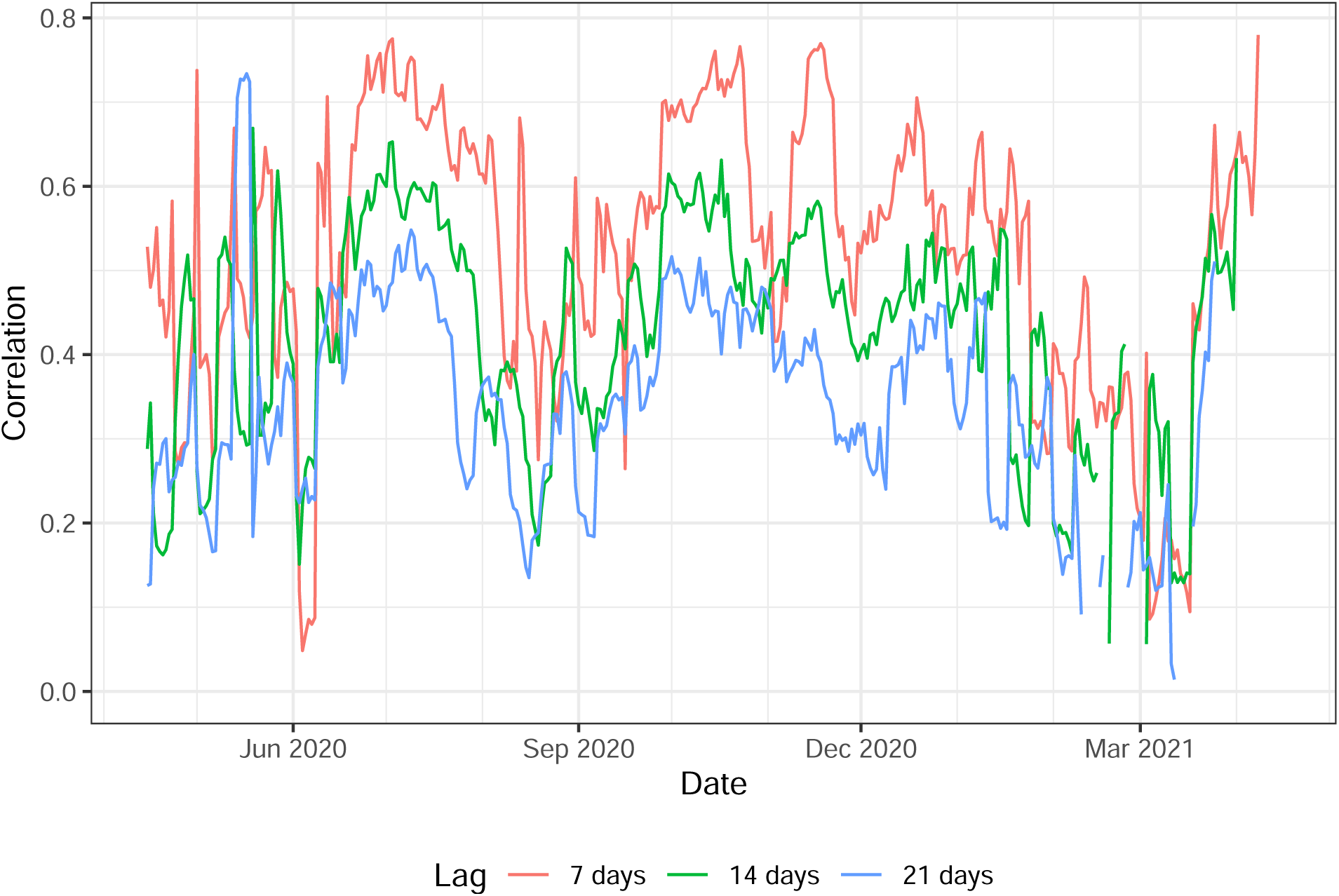
Geo-wise correlations between cases and lagged cases 1, 2, or 3 weeks prior, for all counties in the U.S. Lagged cases are correlated with cases as one might expect, but note the precipitous drop in correlation in February 2021. This matches the correlation drop between other COVIDcast signals and cases during the same time period, supporting the hypothesis that the drop was due to decreased heterogeneity in case rates by county.

**Figure 10:**
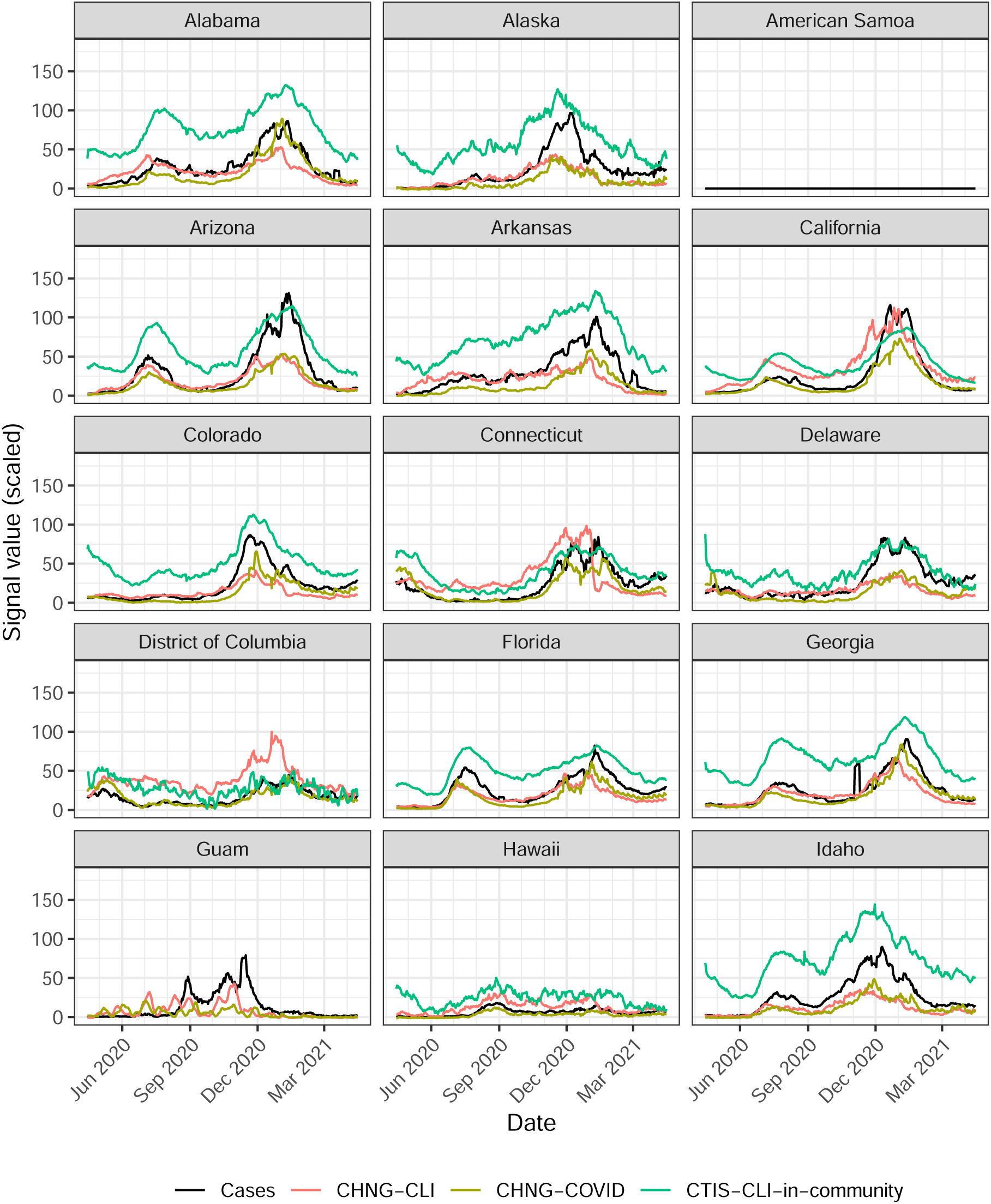
Trends of cases, CHNG-CLI, CHNG-COVID, and CTIS-CLI-in-community for U.S. states and territories. Cases are displayed on the rate scale: counts per 100,000 people. Other signals are scaled to have the same global range across all counties and times. (Part 1 of 4.)

**Figure 11:**
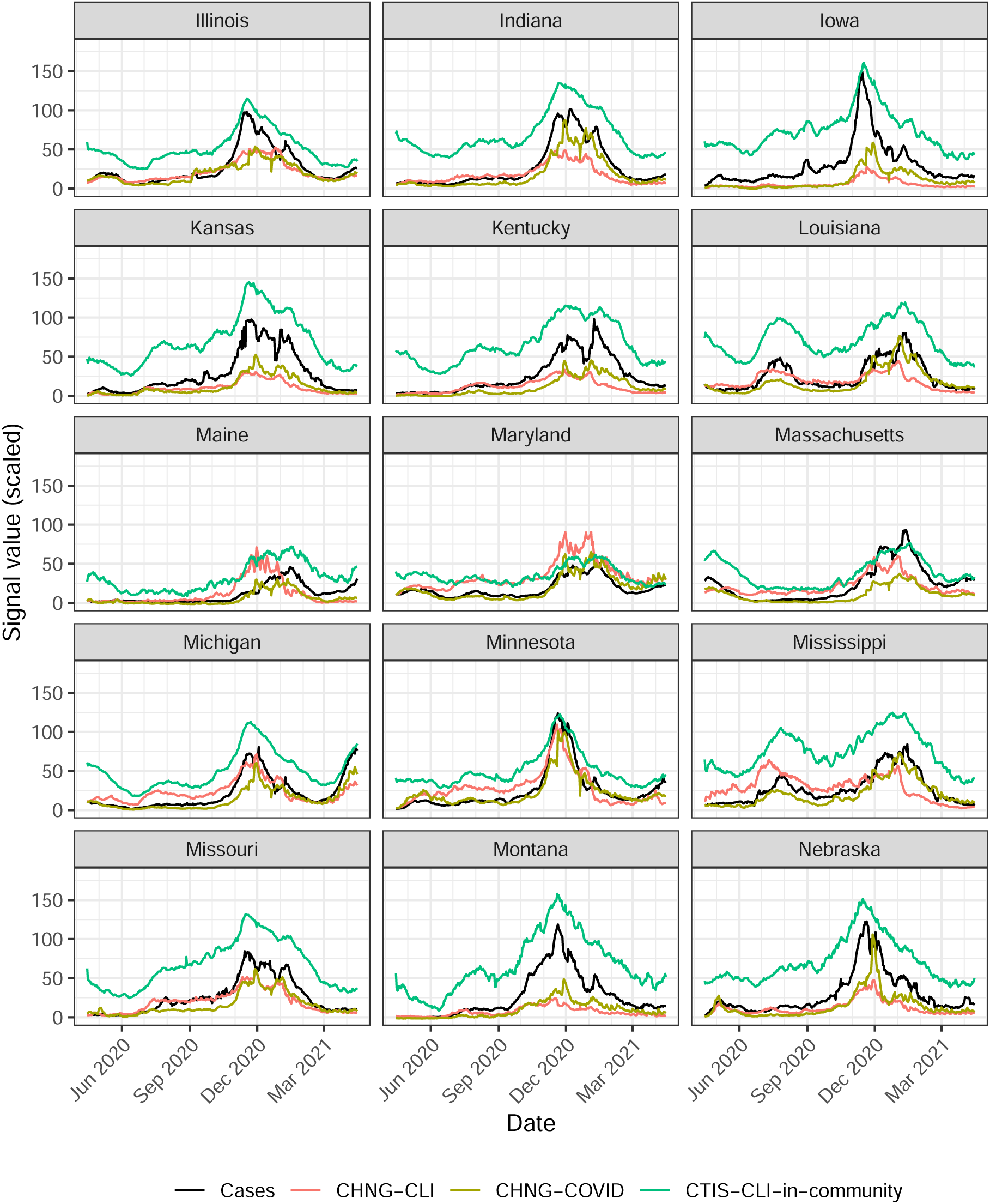
Trends of cases, CHNG-CLI, CHNG-COVID, and CTIS-CLI-in-community for U.S. states and territories. (Part 2 of 4.)

**Figure 12:**
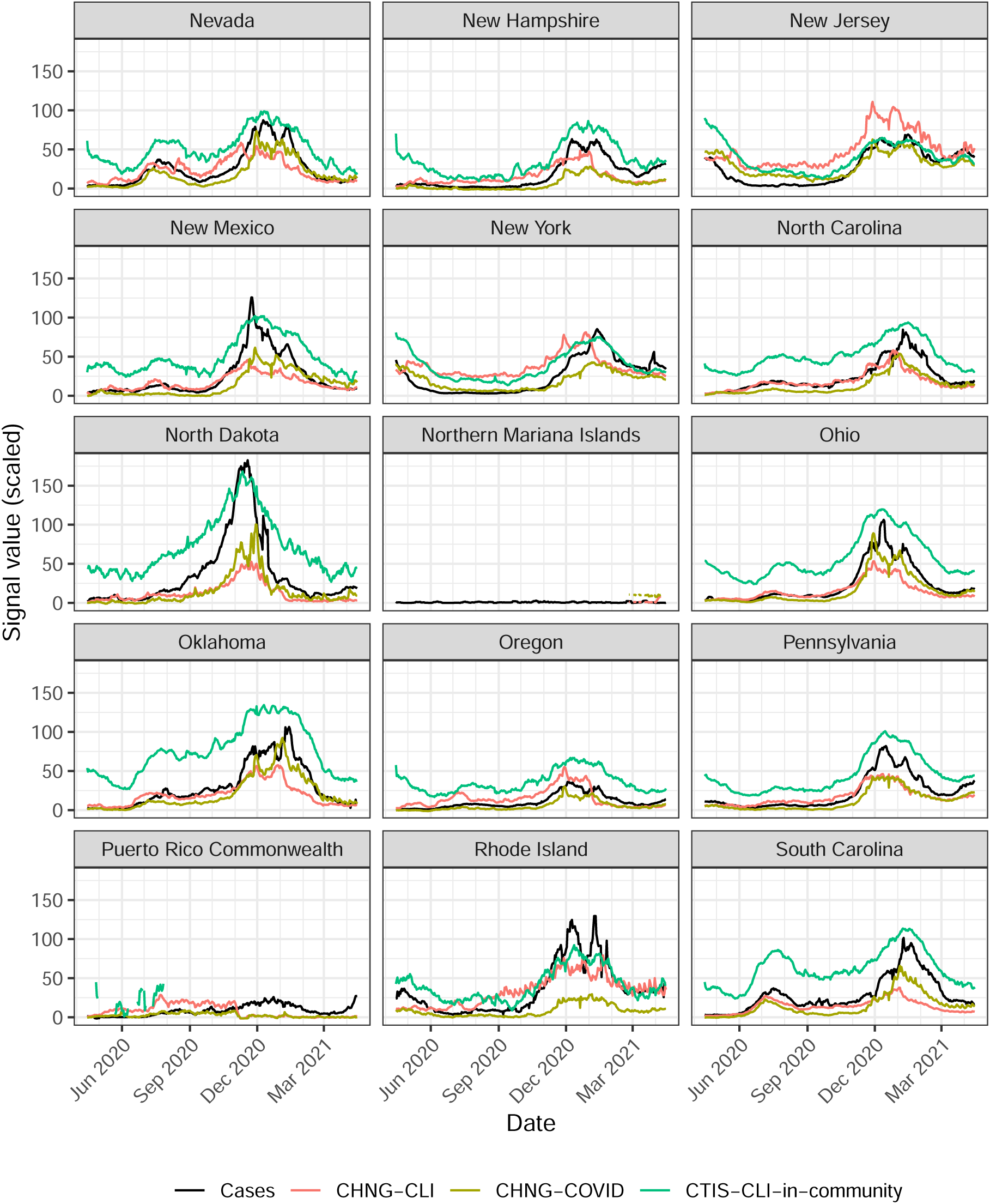
Trends of cases, CHNG-CLI, CHNG-COVID, and CTIS-CLI-in-community for U.S. states and territories. (Part 3 of 4.)

**Figure 13:**
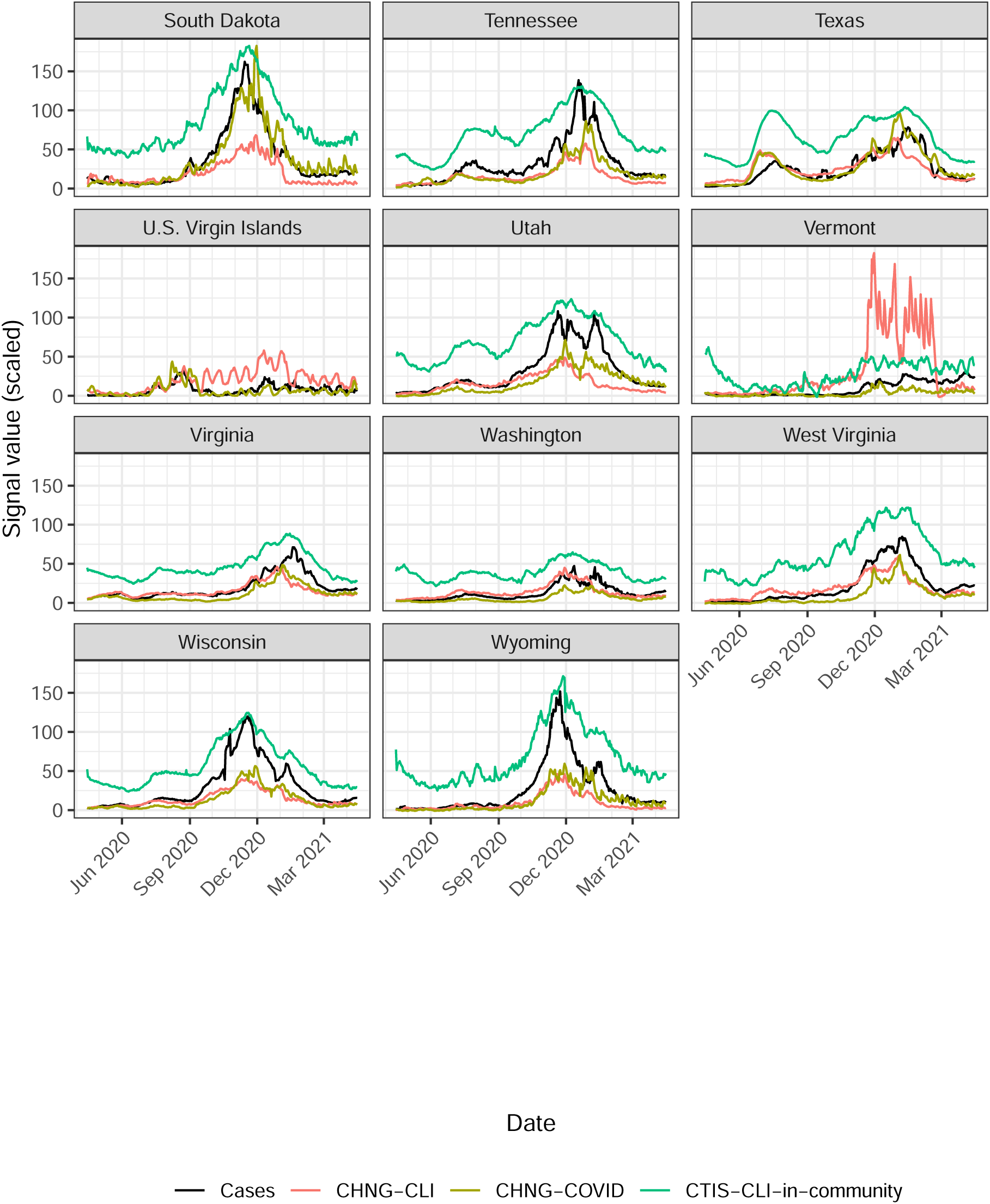
Trends of cases, CHNG-CLI, CHNG-COVID, and CTIS-CLI-in-community for U.S. states and territories. (Part 4 of 4.)

**Figure 14:**
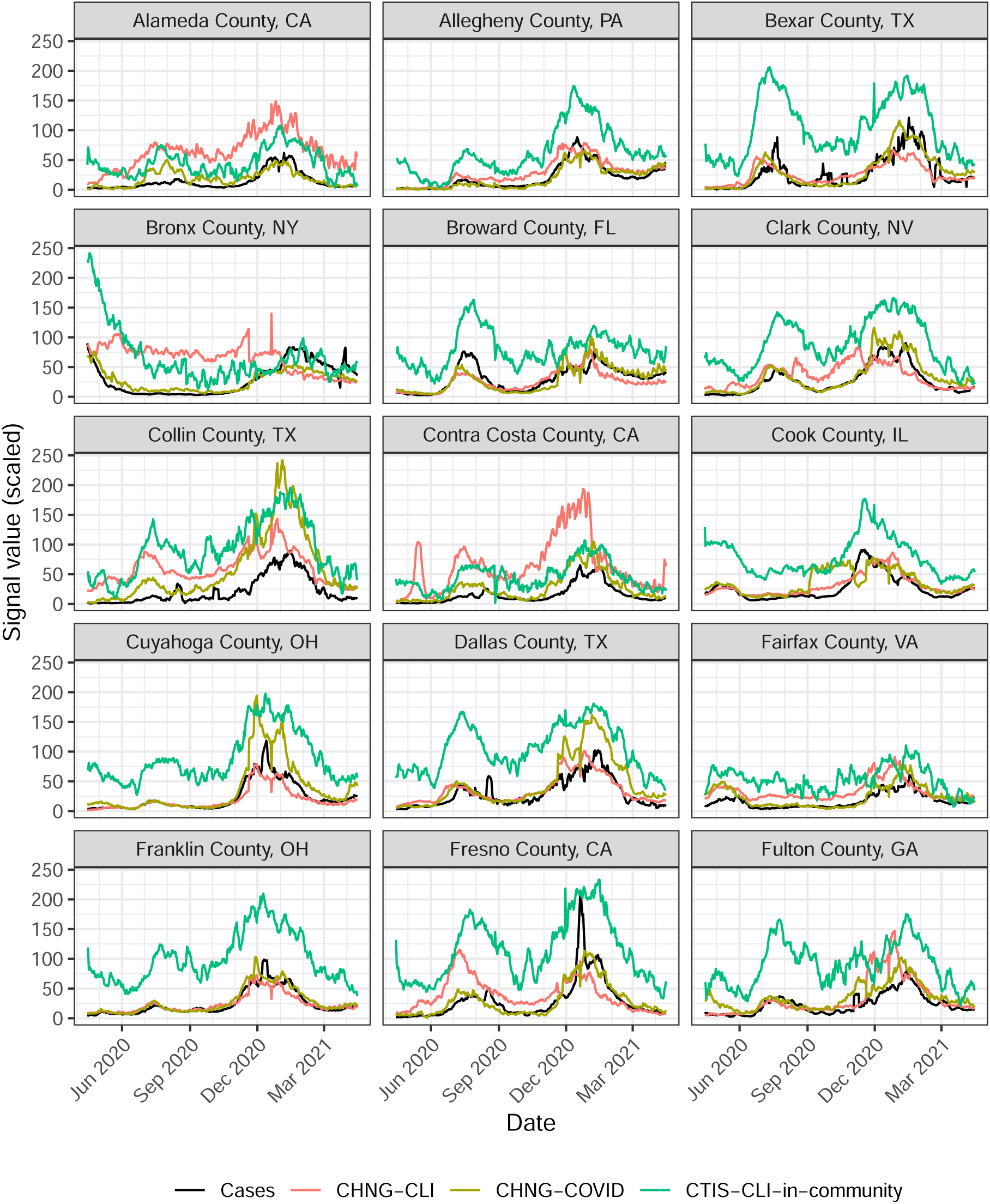
Trends of cases, CHNG-CLI, CHNG-COVID, and CTIS-CLI-in-community for the 50 most populous U.S. counties. Cases are displayed on the rate scale: counts per 100,000 people. Other signals are scaled to have the same global range across all counties and times. (Part 1 of 4.)

**Figure 15:**
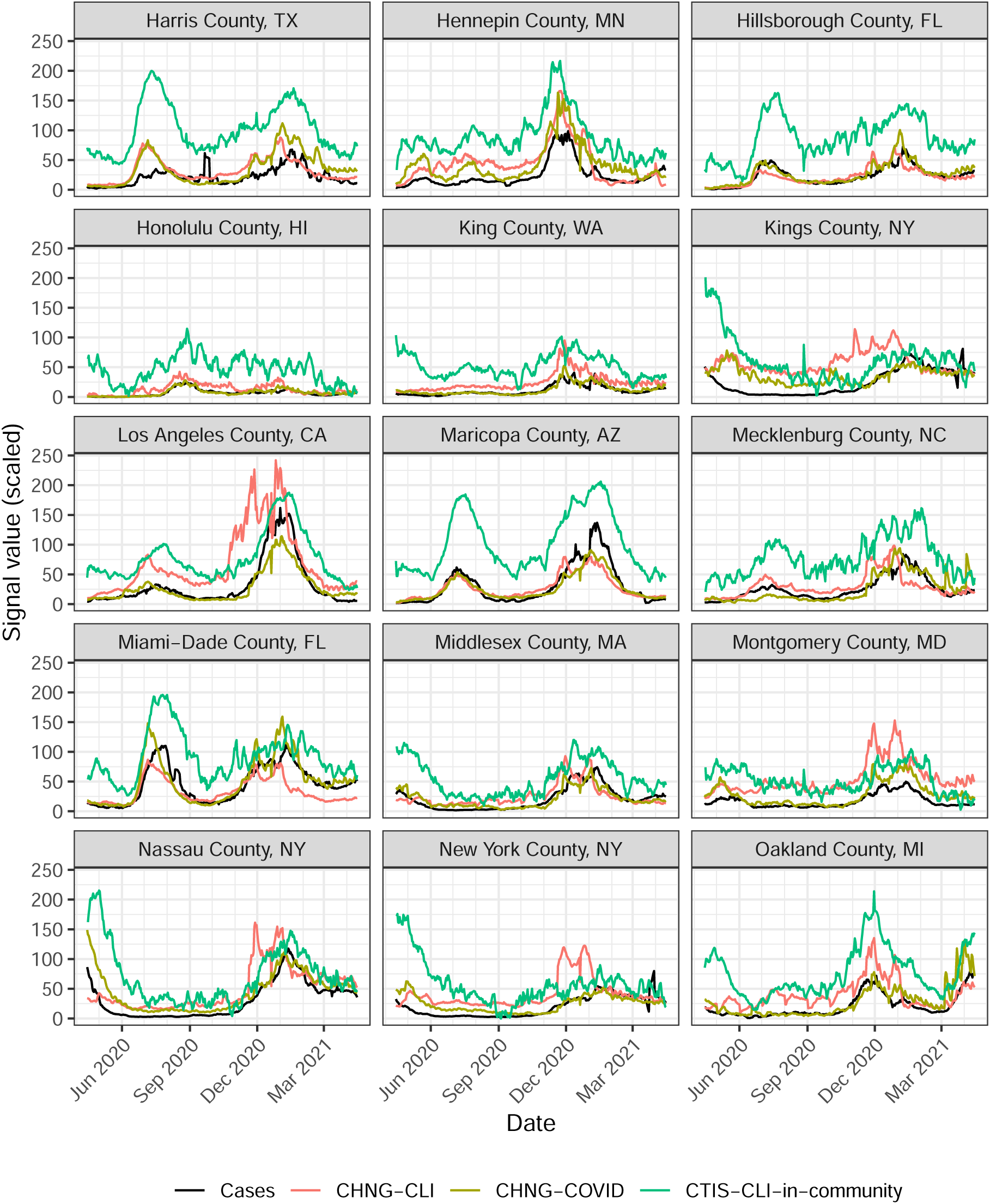
Trends of cases, CHNG-CLI, CHNG-COVID, and CTIS-CLI-in-community for the 50 most populous U.S. counties. (Part 2 of 4.)

**Figure 16:**
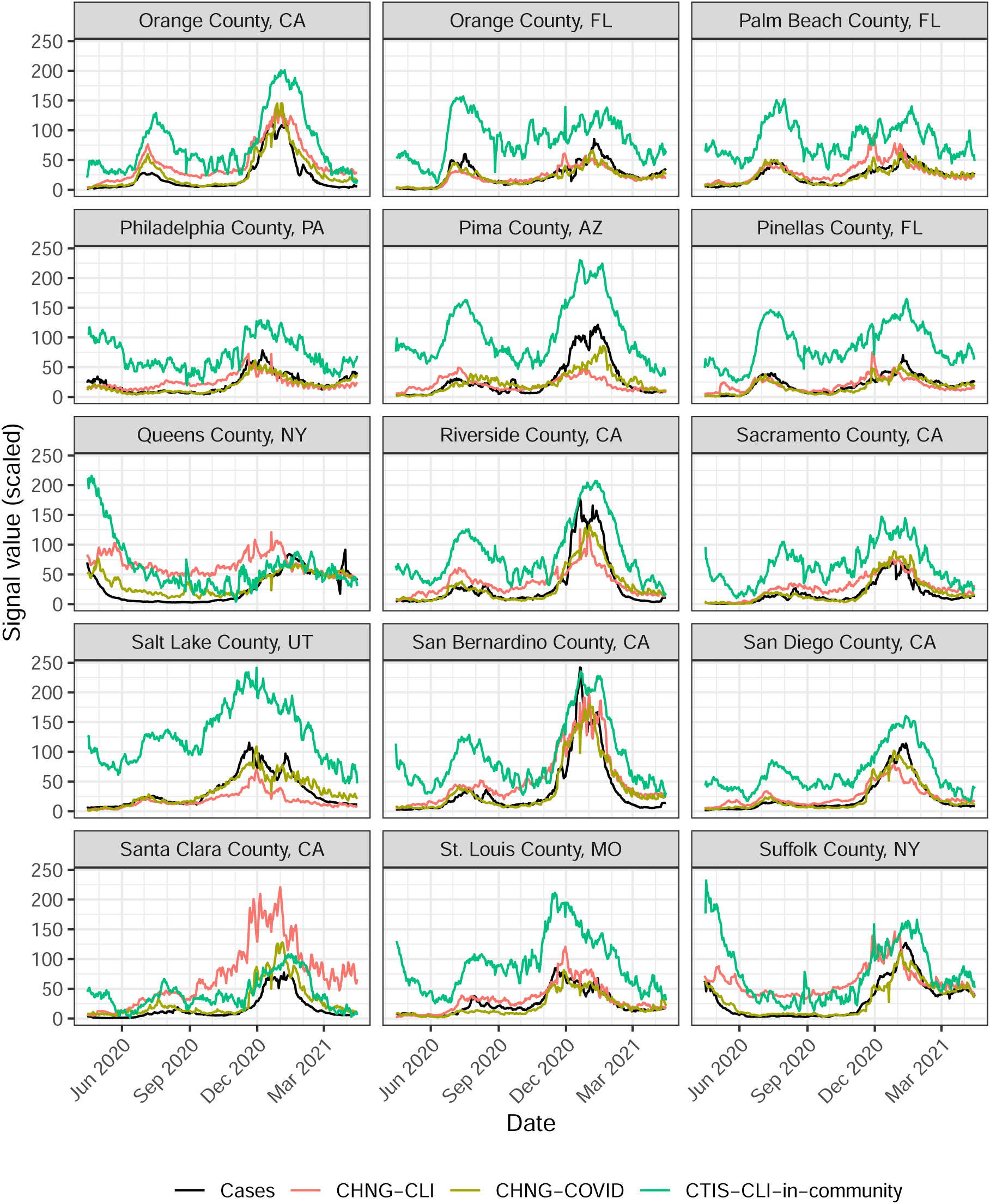
Trends of cases, CHNG-CLI, CHNG-COVID, and CTIS-CLI-in-community for the 50 most populous U.S. counties. (Part 3 of 4.)

**Figure 17:**
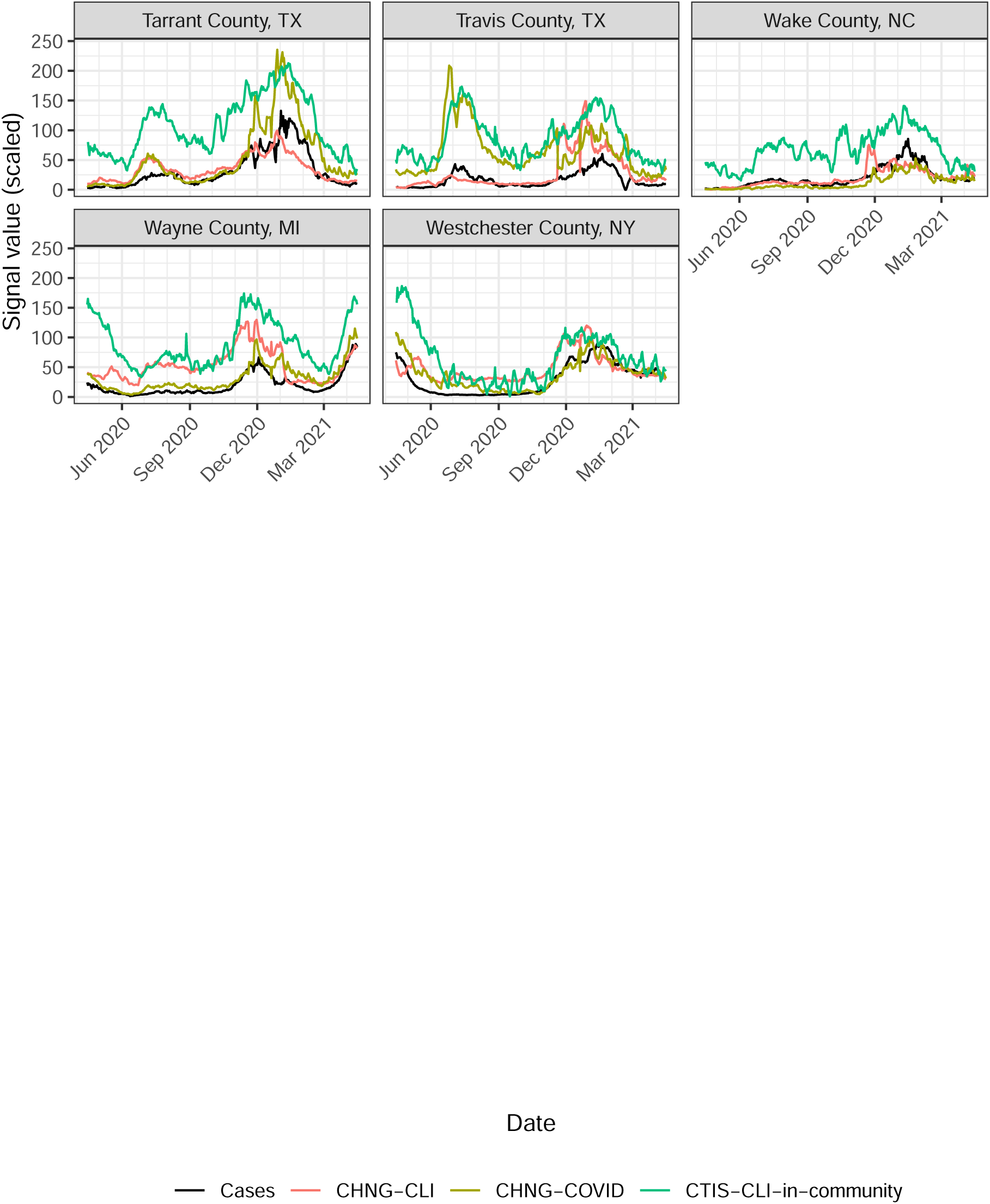
Trends of cases, CHNG-CLI, CHNG-COVID, and CTIS-CLI-in-community for the 50 most populous U.S. counties. (Part 4 of 4.)

**Figure 18:**
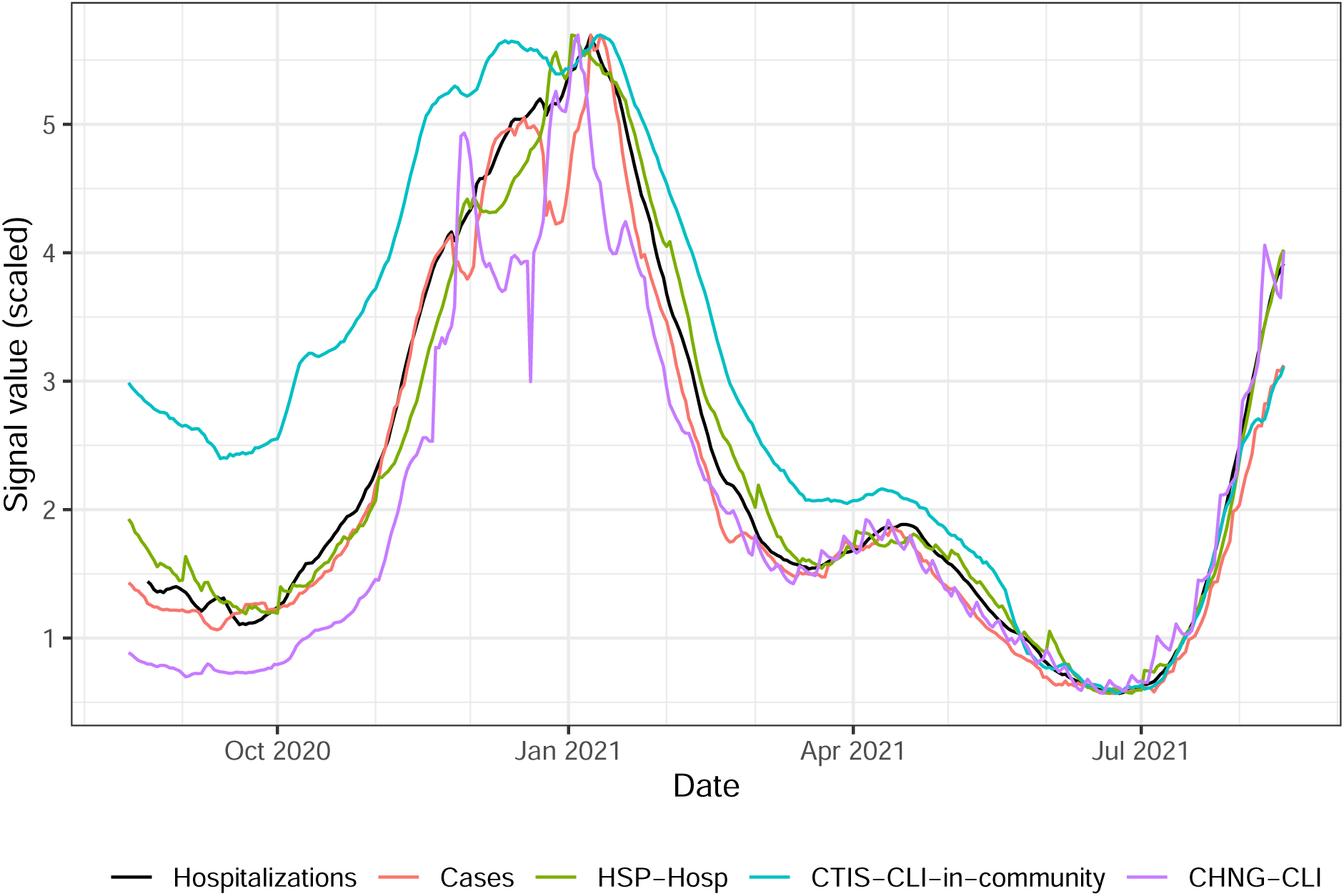
National trends, from August 2020 to August 2021, of HHS-reported confirmed COVID-19 hospital admissions, along with several signals from the COVIDcast API. (HHS data was not consistently reported before August 2020; furthermore, as with cases in the previous trend plots, the HHS data has been smoothed using a 7-day trailing average.) Hospitalizations are displayed on the rate scale: counts per 100,000 people. Other signals are scaled to have the same range. HSP-Hosp is the percentage of new hospital admissions with COVID-associated diagnoses, based on claims data from health system partners (smoothed in time and adjusted for systematic day-of-week effects).

**Figure 19:**
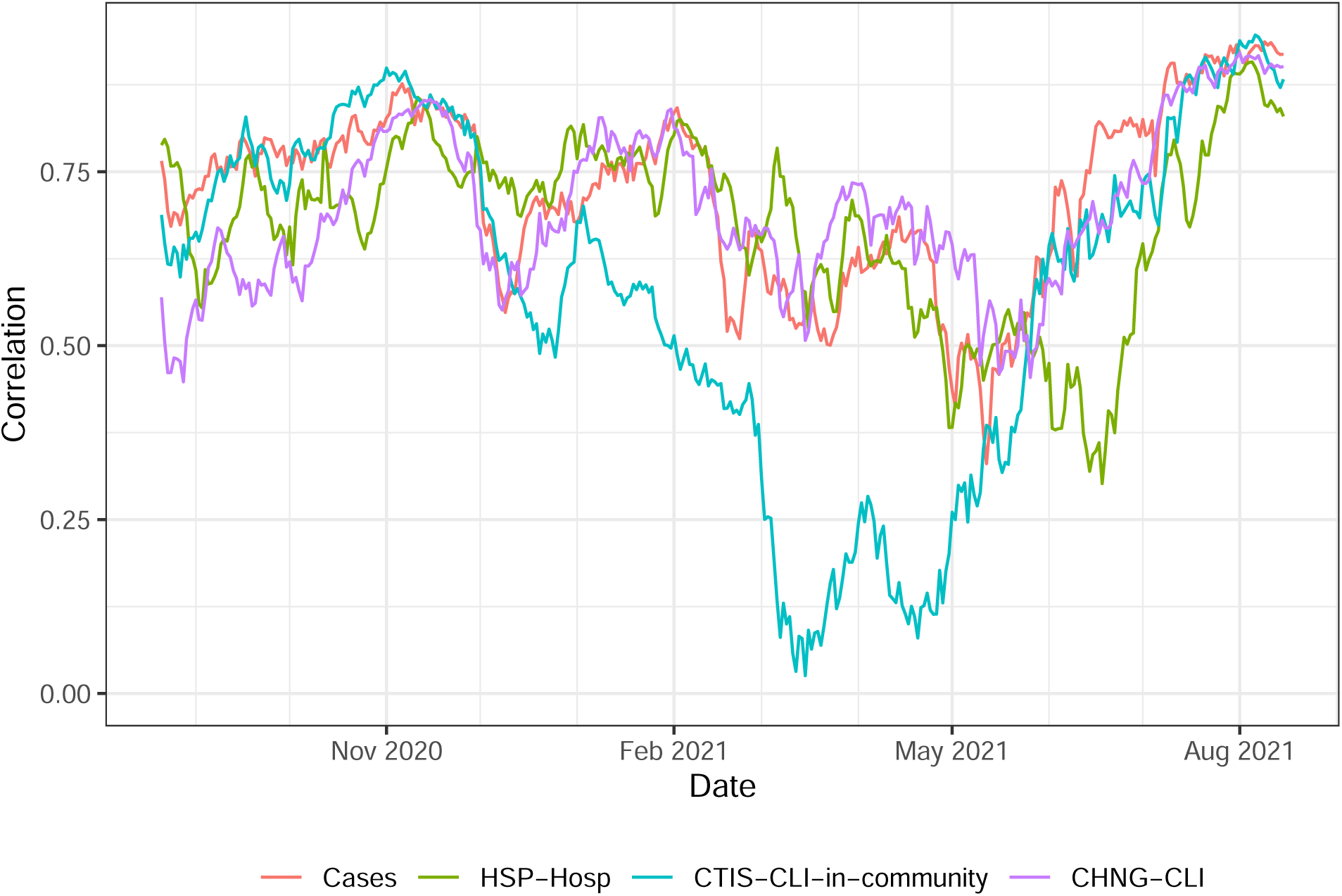
Geo-wise correlations with hospitalization rates derived from HHS data, from August 15, 2020 to August 15, 2021, calculated for all times with sufficient available data within this period, over all state-like jurisdictions for which each signal was reported on at least 50 days during this period, limited to state-day combinations for which all signals are available.

**Figure 20:**
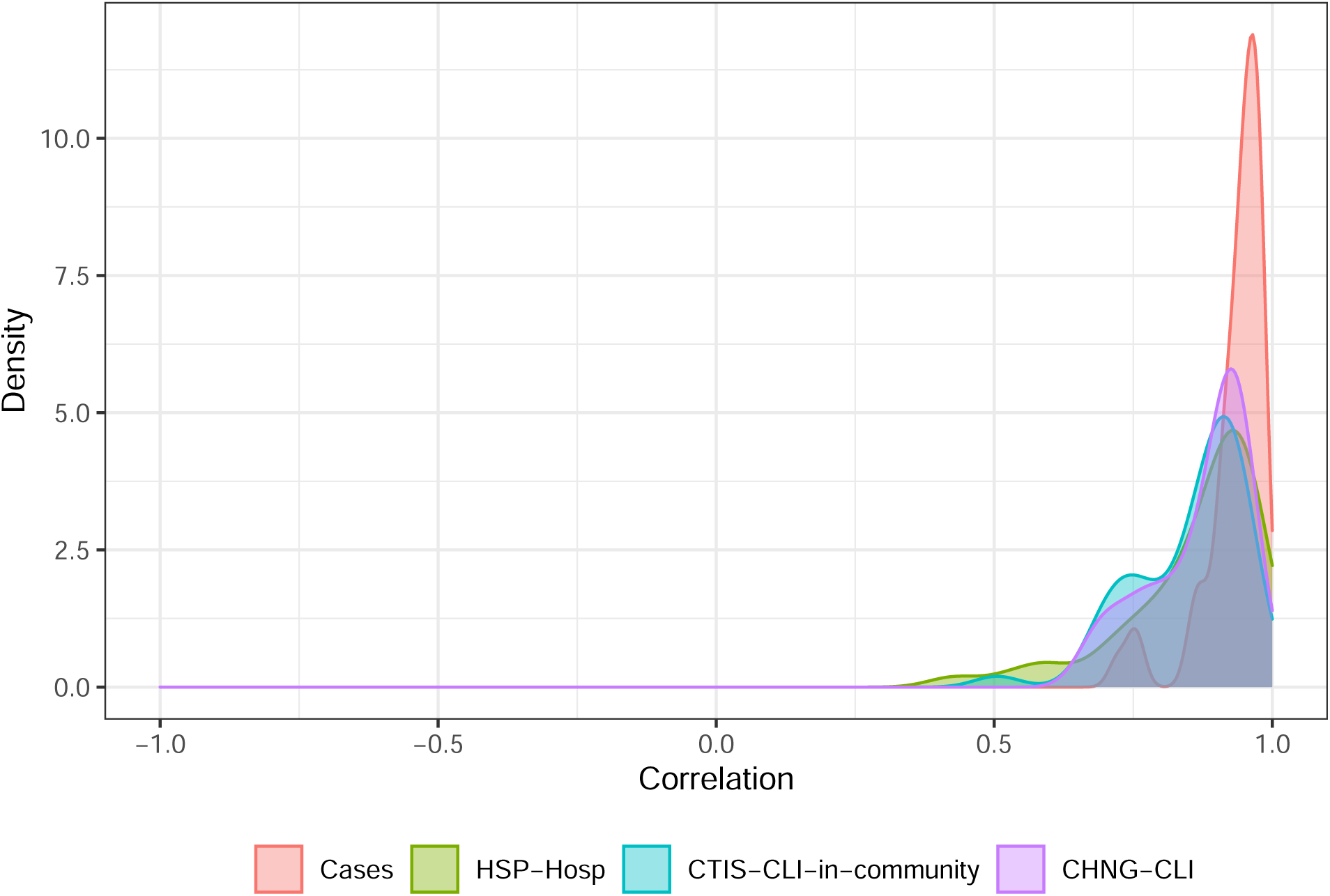
Time-wise correlations with hospitalization rates derived from HHS data, from August 15, 2020 to August 15, 2021, calculated over all state-like jurisdictions for which each signal was reported on at least 50 days during this period, limited to state-day combinations for which all signals are available.

## Notes

### Competing Interest Statement

The authors have declared no competing interest.

### Author Declarations

The Carnegie Mellon University Institutional Review Board reviewed the surveys as STUDY2020_00000162, and determined that the use of deidentified medical claims and test results was not human subjects research. All other data is from publicly available sources.

### Summary of Updates

Expanded supplementary information; clarified limitations; new analyses in Results section.

